# Thyroid function abnormalities in COVID-19 patients

**DOI:** 10.1101/2020.06.15.20130807

**Authors:** Weibin Wang, Xingyun Su, Yongfeng Ding, Weina Fan, Junwei Su, Zhendong Chen, Hong Zhao, Kaijin Xu, Qin Ni, Xiaowei Xu, Yunqing Qiu, Lisong Teng

**Affiliations:** Department of Surgical Oncology, The First Affiliated Hospital, Zhejiang University School of Medicine, Hangzhou, Zhejiang, China; Department of Medical Oncology, The First Affiliated Hospital, Zhejiang University School of Medicine, Hangzhou, Zhejiang, China; Department of Intensive Care Unit (ICU), The First Affiliated Hospital, Zhejiang University School of Medicine, Hangzhou, Zhejiang, China; State Key Laboratory for Diagnosis and Treatment of Infectious Diseases, National Clinical Research Center for Infectious Diseases, Collaborative Innovation Center for Diagnosis and Treatment of Infectious Diseases, Department of Infectious Diseases, The First Affiliated Hospital, College of Medicine, Zhejiang University, Hangzhou, Zhejiang, China

**Keywords:** COVID-19, thyroid function, abnormality, thyroid stimulating hormone, pathogenesis

## Abstract

**Background:** The novel coronavirus COVID-19, has caused a worldwide pandemic, impairing several human organs and systems. Whether COVID-19 affects human thyroid function remains unknown.

**Methods:** 84 hospitalized COVID-19 patients in the First Affiliated Hospital, Zhejiang University School of Medicine (Hangzhou, China) were respectively enrolled in this study. In addition, 91 other patients with pneumonia and 807 healthy subjects were included as controls.

**Findings:** We found that the levels of TT3 and TSH were lower in COVID-19 patients than control groups (p<0·001). Within the group of COVID-19 patients, 61.9% patients (52/84) presented with thyroid function abnormalities. We found a larger proportion of patients in severe condition exhibited thyroid dysfunction than mild/moderate cases (74·6% vs. 23·8%, p < 0·001). Patients with thyroid dysfunction tended to have increased interval time for negative conversion of viral nucleic acid (14·1 ± 9·4 vs. 10·6 ± 8·3 days, p = 0·088). To note, thyroid dysfunction was also associated with decreased lymphocytes (p < 0·001) and increased CRP (p = 0·002). In 7 patients with dynamic changes of thyroid function, we observed the levels of TT3 and TSH gradually increased and reached normal range without thyroid hormone replacement at Day 30 post-admission. The correlation between TT3 and TSH level seemed to be positive rather than negative in the early stage, and gradually turned to be negatively related over time.

**Interpretations:** Thyroid function abnormalities are common in COVID-19 patients, especially in severe cases. This might be caused by virus attack and damage to the thyroid-pituitary axis. Therefore, more attention should be paid to thyroid function during treatment of COVID-19, and close follow-up is also needed after discharge.

**Funding:** This study was supported by Zhejiang Provincial Science and technology department key R & D plan emergency project (No. 2020c03123-8).

## Introduction

The outbreak of coronavirus disease 2019 (COVID-19), caused by severe acute respiratory syndrome coronavirus 2 (SARS-CoV-2), has rapidly spread worldwide and led to the declaration of Public Health Emergency of International Concern by the World Health Organization (WHO) ^1-3^. As of June 14, 2020, a total of 7, 844, 978 cases have been confirmed worldwide, among them, 428, 045 people have died of COVID-19. Patients infected with COVID-19 display mainly symptoms similar with pneumonia such as fever, fatigue, cough, shortness of breath ^4,5^. And many patients have symptoms outside of the respiratory system including poor appetite, diarrhea, nausea, vomiting, palmus, and chest distress ^4,5^. The main management of COVID-19 infection is supportive, and acute respiratory distress syndrome (ARDS) induced respiratory failure is the leading cause of mortality ^4,6,7^.

Severe and complex effects on several human organs and systems including respiratory, immune, digestive, circulatory, hepatic, renal, and hematological systems have been reported in COVID-19 patients ^4-8^. Whether COVID-19 affects human thyroid function remains unknown. Thyroid dysfunction was identified in severe acute respiratory syndrome (SARS) patients, which was caused by another strain of coronavirus. Recently, Brancatella et al. reported the first case of subacute thyroiditis after SARS-CoV-2 infection ^9^. Both indicate that the thyroid gland may also be a target organ of SARS-CoV-2.

In the present study, 84 hospitalized COVID-19 patients in the First Affiliated Hospital, Zhejiang University School of Medicine (Hangzhou, China) were respectively enrolled. In addition, 91 other patients with pneumonia and 807 healthy subjects were included as controls. The thyroid function in COVID-19, pneumonia patients and healthy subjects was compared, and its relationship with disease severity, interval time for negative conversion of viral nucleic acid, auto-antibodies, leukocytes, inflammatory biomarkers and cytokines were also investigated to uncover the underlying clinical value of thyroid dysfunction. Furthermore, the dynamic changes of thyroid function during recovery were analyzed to depict the development of thyroid dysfunction induced by SARS-CoV-2.

## Materials and Methods

### Participants

There were 96 hospitalized patients definitely diagnosed as COVID-19 according to WHO interim guidance ^10^ in the First Affiliated Hospital, Zhejiang University School of Medicine (Hangzhou, China). All the COVID-19 patients admitted to our hospital in January 22^st^ to March 16^st^, 2020. Among them, 85 patients had detected thyroid function after admission. One patient was excluded due to pregnancy. In total, 84 COVID-19 patients were finally enrolled in this retrospective study. Meanwhile, 91 non-COVID-19 pneumonia patients and 807 healthy subjects were also included as controls. These 91 non-COVID-19 cases were matched age, gender, and clinical classification with COVID-19 patients, who were infected by bacteria, fungus, and virus at the same period.

Clinical classification of COVID-19 was according to the Handbook of COVID-19 Prevention and Treatment ^11^, and severe cases were judged when met any of the following criteria: 1) respiratory rate over 30 breaths / min; 2) oxygen saturation ≤ 93% at a rest state; 3) arterial partial pressure of oxygen (PaO2) / oxygen concentration (FiO2) ≤ 300 mmHg. Besides, patients with > 50% lesions progression within 24 to 48 hours in lung imaging should be regarded as severe cases. All the cases were obtained with informed consent. The research was approved by the Ethics Committee of the First Affiliated Hospital, Zhejiang University School of Medicine.

### Data Collection and Procedures

We compared age, gender, and thyroid function including triiodothyronine (TT3), thyroxine (TT4), free triiodothyronine (FT3), free thyroxine (FT4), and thyroid stimulating hormone (TSH) among COVID-19, non-COVID-19 pneumonia patients and healthy subjects. The association between thyroid function and disease severity, inflammatory biomarker (leukocytes, C-reactive protein, procalcitonin), inflammatory cytokines (interleukin-6, interleukin-10, tumor necrosis factor-α, interferon-γ), auto-antibodies (thyroglobulin antibody, thyroid peroxidase antibody), and interval time for negative conversion of viral nucleic acid were analyzed. We also studied the dynamic changes of thyroid function during patients’ recovery period.

### Statistical Analysis

Statistical analysis was conducted by SPSS (version 21·0) (SPSS Inc., Chicago, IL, USA) and R language (Version 3·6·3). Pearson chi-square test and analysis of variance (ANOVA) were used to analyze the characteristics and thyroid function among COVID-19, non-COVID-19 pneumonia patients and healthy subjects, and to establish the factors associated with dysfunction of thyroid. Pearson correlation was performed in the correlation analysis. In addition, a polynomial regression curve was fitted between time after hospitalization and TSH or TT3 levels. For all analyses, p < 0·05 was regarded as statistically significant.

## Results

### Characteristics of participants

A total of 84 hospitalized patients diagnosed as COVID-19 were recruited in our study. All 84 COVID-19 patients had record of TT3, TT4, and TSH at admission, with 22 patients having complete record of thyroid function including FT3 and FT4. We analyzed the association between FT3 and TT3 and found that FT3 was positively correlated with TT3 (p < 0·001) (Figure S1) indicating that changes of FT3 can be reflected TT3 to a certain extent.

Among the COVID-19 patients, 63 patients were clinically classified as severe. The mean age of COVID-19 patients was 57·3 ± 14·5 years old, and 63% (53/84) were male, no significance was found when compared with non-COVID-19 pneumonia patients or healthy subjects (Table 1). Based on quantification of thyroid hormones, a total of 52 (52/84, 61·9%) COVID-19 patients were defined as thyroid dysfunction. Among them, only two patients had increased TT4 and decreased TSH levels, while the others had lower than normal range on either TT4, TT3, or TSH.

**Table 1.** Comparison of clinical features among COVID-19, non-COVID-19 pneumonia patients and healthy subjects

| Characteristics | COVID-19 patients<br>(N=84)<br>Mean ± SD or n (%) | Non-COVID-19 pneumonia patients<br>(N=91)<br>Mean ± SD or n (%) | Healthy subjects<br>(N=807)<br>Mean ± SD or n (%) | *p* value | | | | |
| --- | --- | --- | --- | --- | --- | --- | --- | --- |
|  |  |  |  | Among groups | COVID-19 vs. Non-COVID-19 |  | COVID-19 vs. Healthy subjects |  |
| Mean age (yrs) | 57.3 ± 14.5 | 60.1 ± 16.7 | 57.7 ± 13.0 | 0.248 | 0.236 | | 0.782 | |
| Gender |  |  |  |  |  |  |  |  |
| Male | 53 (63.1%) | 60 (65.9%) | 474 (58.7%) | 0.336 | 0.695 |  | 0.439 |  |
| Female | 31 (36.9%) | 31 (34.1%) | 333 (41.3%) |  |  |  |  |  |
| Clinical classifications |  |  |  |  |  |  |  |  |
| Mild and moderate | 21 (25.0%) | 24 (26.4%) | - | - | 0.835 |  | - |  |
| Severe and critical | 63 (75.0%) | 67 (73.6%) |  |  |  |  |  |  |
| Thyroid Function |  |  |  |  |  |  |  |  |
| TT4 (nmol/L) | 99.04 ± 25.96 | 82.28 ± 26.47 | 97.25 ± 17.13 | **0.000** | **0.000** | | 0.391 | |
| TT3 (nmol/L) | 1.02 ± 0.32 | 0.92 ± 0.38 | 1.59 ± 0.24 | **0.000** | 0.059 | | **0.000** | |
| TSH (mIU/L) | 0.62 ± 0.62 | 1.07 ± 0.94 | 1.55 ± 0.94 | **0.000** | **0.000** | | **0.000** | |

TT4, total thyroxine or tetraiodothyronine, normal range 62.68-150.84 nmol/L; TT3, total triiodothyronine normal range 0.89-2.44 nmol/L; TSH, thyroid-stimulating hormone, normal range 0.35-4.94 nmol/L.
*p* value in bold was regarded as statistically significant.

### Thyroid dysfunction in COVID-19 patients

We analyzed the levels of TT4, TT3, and TSH on admission in COVID-19, non-COVID-19 pneumonia patients and healthy subjects. The level of TT3 in COVID-19 patients (1·02 ± 0·32 nmol/L) was lower than that in healthy subjects (1·59 ± 0·24 nmol/L) (p < 0·001), but similar with non-COVID-19 patients (0·92 ± 0·38 nmol/L) (p=0·59) (Table 1). There was no obvious difference for TT4 between COVID-19 patients and healthy subjects (p = 0·391) (Table 1). The thyroid gland is an endocrine organ negatively regulated by TSH which is secreted by the adenohypophysis. However, in COVID-19 cases, TSH level surprisingly decreased by a lower extent (0·62 ± 0·62 mIU/L) as compared to non-COVID-19 patients (1·07 ± 0·94 mIU/L) and healthy subjects (1·55 ± 0·94 mIU/L) (p < 0.001) (Table 1). Correlation analysis showed that the levels of TT3 and TSH were positively correlated, instead of negatively related, in COVID-19 patients (R = 0·575, p < 0·001) (Figure 1).

**Figure 1.**
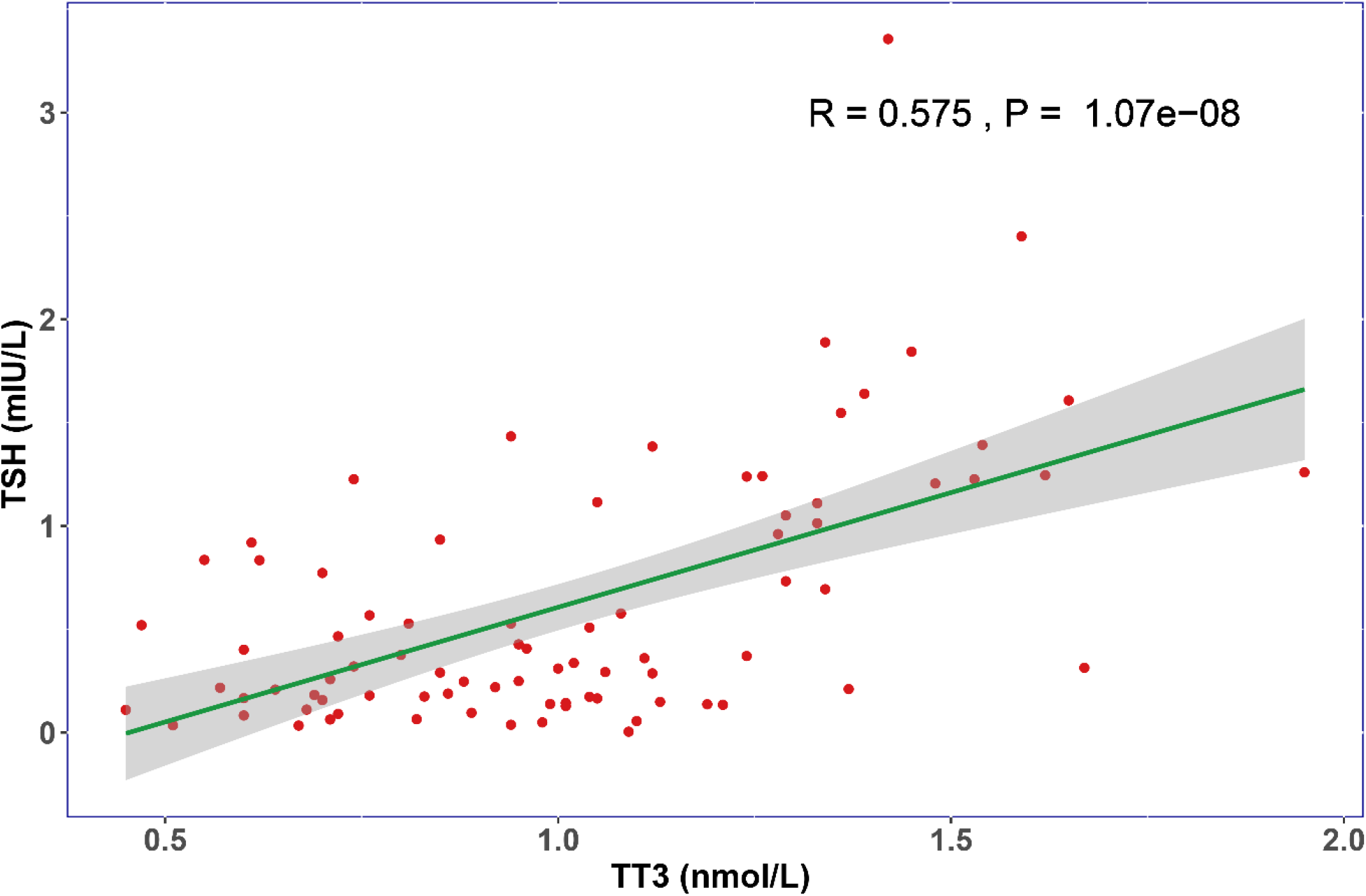
The relationship of TT3 and TSH levels on admission. The TT3 and TSH levels in COVID-19 patients are positively correlated (R=0.575, P<0.001)

### Analysis of clinical values for thyroid dysfunction in COVID-19 patients

COVID-19 patients were further divided into thyroid dysfunction group and normal group according to TT3, TT4, and TSH levels. No obvious difference was found in age and sex between two groups (Table 2). Abnormal thyroid dysfunction was more common in severe cases (74·6%) than mild/moderate cases (23·8%) (p < 0·001) (Table 2). Interestingly, thyroid dysfunction tended to be associated with longer interval time for negative conversion of viral nucleic acid (14·13 ± 9·39 vs. 10·56 ± 8·29 days, p = 0·088). Besides, increased levels of leucocytes (p < 0·001), neutrophils (p < 0·001), CRP (p = 0·002) and PCT (p = 0·054), and decreased level of lymphocytes (p < 0·001) were more prevalent in thyroid dysfunction group. Meanwhile, we did not find any significance in auto-antibodies (thyroglobulin antibody, thyroid peroxidase antibody) and inflammatory cytokines (IL-6, IL-10, TNF-α, IFN-γ) (Table 2).

**Table 2.**
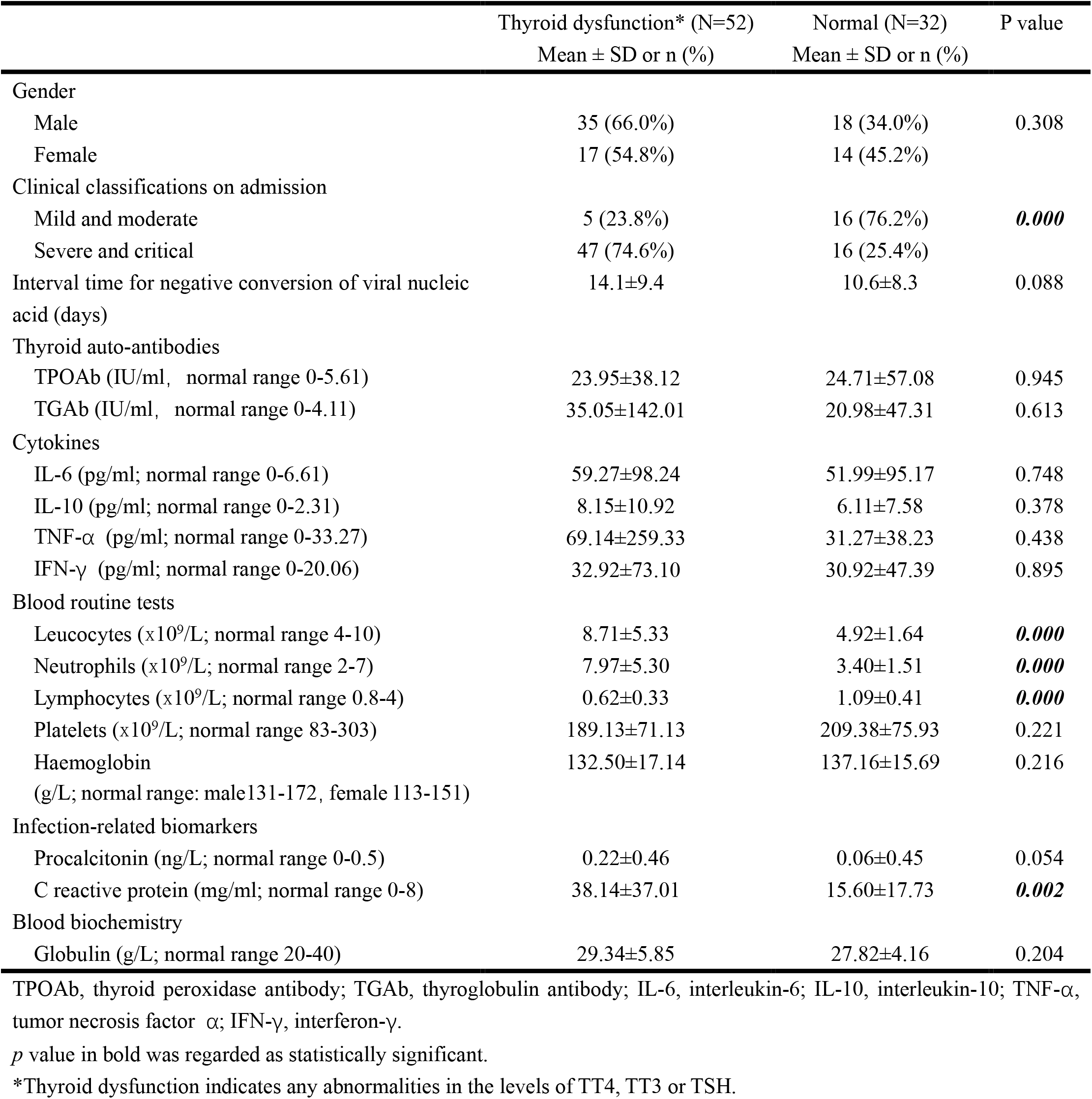
Clinical characteristics and selected laboratory abnormalities of COVID-19 patients with and without thyroid dysfunction

| | Thyroid dysfunction* (N=52)<br>Mean ± SD or n (%) | Normal (N=32)<br>Mean ± SD or n (%) | P value |
| --- | --- | --- | --- |
| Gender |  |  |  |
| Male | 35 (66.0%) | 18 (34.0%) | 0.308 |
| Female | 17 (54.8%) | 14 (45.2%) |  |
| Clinical classifications on admission |  |  |  |
| Mild and moderate | 5 (23.8%) | 16 (76.2%) | **0.000** |
| Severe and critical | 47 (74.6%) | 16 (25.4%) |  |
| Interval time for negative conversion of viral nucleic acid (days) | 14.1±9.4 | 10.6±8.3 | 0.088 |
| Thyroid auto-antibodies |  |  |  |
| TPOAb (IU/ml, normal range 0-5.61) | 23.95±38.12 | 24.71±57.08 | 0.945 |
| TGAb (IU/ml, normal range 0-4.11) | 35.05±142.01 | 20.98±47.31 | 0.613 |
| Cytokines |  |  |  |
| IL-6 (pg/ml; normal range 0-6.61) | 59.27±98.24 | 51.99±95.17 | 0.748 |
| IL-10 (pg/ml; normal range 0-2.31) | 8.15±10.92 | 6.11±7.58 | 0.378 |
| TNF-α (pg/ml; normal range 0-33.27) | 69.14±259.33 | 31.27±38.23 | 0.438 |
| IFN-γ (pg/ml; normal range 0-20.06) | 32.92±73.10 | 30.92±47.39 | 0.895 |
| Blood routine tests |  |  |  |
| Leucocytes ($\times 10^9$/L; normal range 4-10) | 8.71±5.33 | 4.92±1.64 | **0.000** |
| Neutrophils ($\times 10^9$/L; normal range 2-7) | 7.97±5.30 | 3.40±1.51 | **0.000** |
| Lymphocytes ($\times 10^9$/L; normal range 0.8-4) | 0.62±0.33 | 1.09±0.41 | **0.000** |
| Platelets ($\times 10^9$/L; normal range 83-303) | 189.13±71.13 | 209.38±75.93 | 0.221 |
| Haemoglobin (g/L; normal range: male 131-172, female 113-151) | 132.50±17.14 | 137.16±15.69 | 0.216 |
| Infection-related biomarkers |  |  |  |
| Procalcitonin (ng/L; normal range 0-0.5) | 0.22±0.46 | 0.06±0.45 | 0.054 |
| C reactive protein (mg/ml; normal range 0-8) | 38.14±37.01 | 15.60±17.73 | **0.002** |
| Blood biochemistry |  |  |  |
| Globulin (g/L; normal range 20-40) | 29.34±5.85 | 27.82±4.16 | 0.204 |

TPOAb, thyroid peroxidase antibody; TGAb, thyroglobulin antibody; IL-6, interleukin-6; IL-10, interleukin-10; TNF-α , tumor necrosis factor $\alpha$ ; IFN-γ , interferon-γ .
*p* value in bold was regarded as statistically significant.
\*Thyroid dysfunction indicates any abnormalities in the levels of TT4, TT3 or TSH.

### The development of thyroid dysfunction in COVID-19 patients

In order to illustrate the development of thyroid dysfunction induced by SARS-CoV-2, we analyzed 7 patients with records of thyroid function during their recovery period. All 7 patients had lower than normal range of TSH levels on admission, and none of them was treated by glucocorticoid or thyroxine. We observed that the levels of TT3 and TSH gradually increased during 2 months after hospitalization (Figure 2). At Day 30 post-admission, all the patients’ TT3 and TSH levels recovered without thyroid hormone replacement (Figure 2). We further found TT3 and TSH levels seem to be altered from positive correlation to negative over time, indicating a recovery of pituitary-thyroid axis. However, given limited patient number, the p values did not achieve significance (Figure 3).

**Figure 2.**
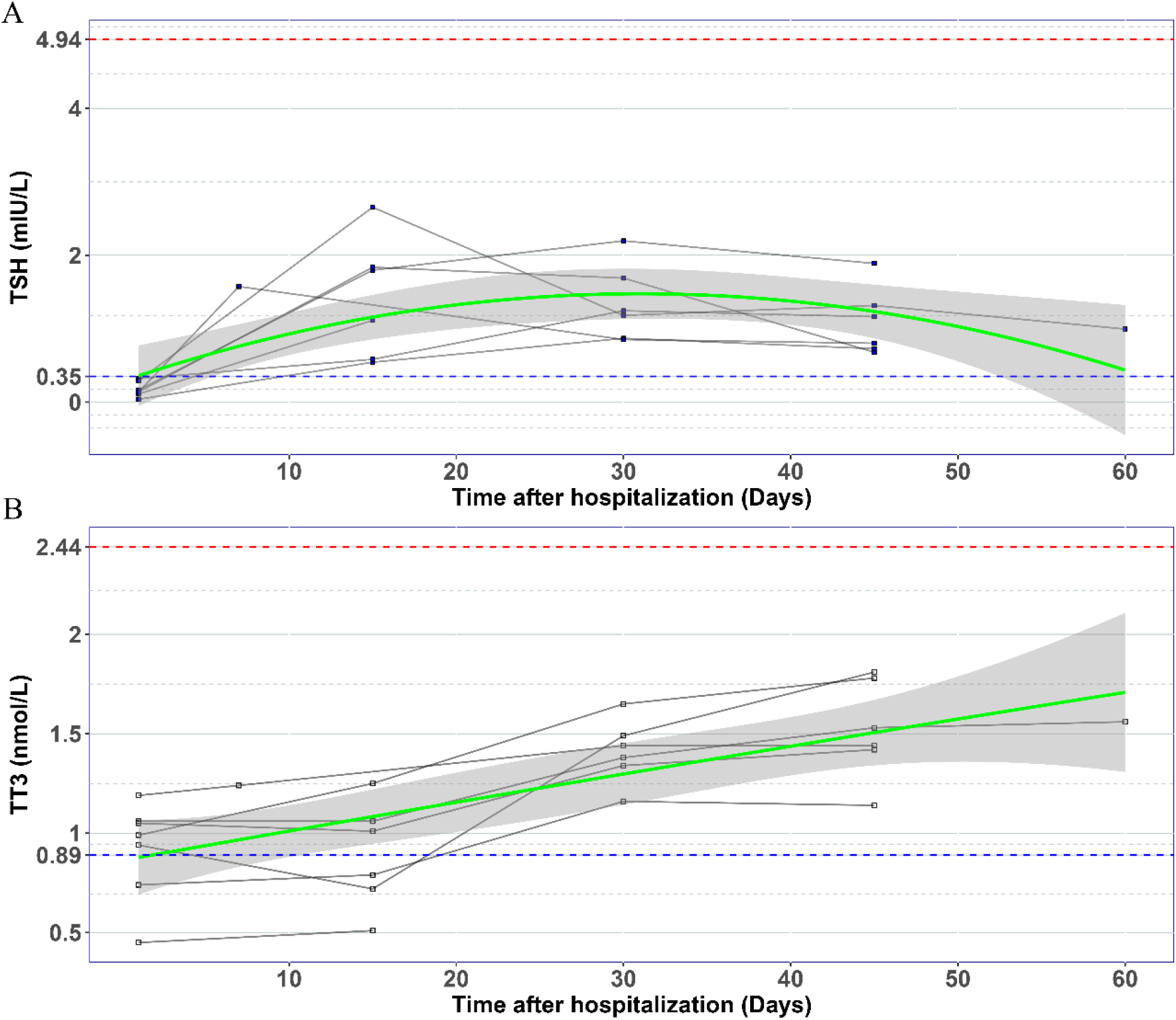
The changes of TSH (A) and TT3 (B) levels during hospitalization in COVID-19 patients with abnormal TSH level on admission. Every polyline represents the variation trend of TSH or TT3 level of one patient. Dashed blue lines, the lower limit of normal TSH (0.35 mIU/L) and TT3 (0.89 nmol/L) value; Dashed red lines, the upper limit of normal TSH (4.94 mIU/L) and TT3 (2.44 nmol/L) value; Green curves represent the fitting of data; Grey shaded areas, depict the 95% confidence band for the fitted curve.

**Figure 3.**
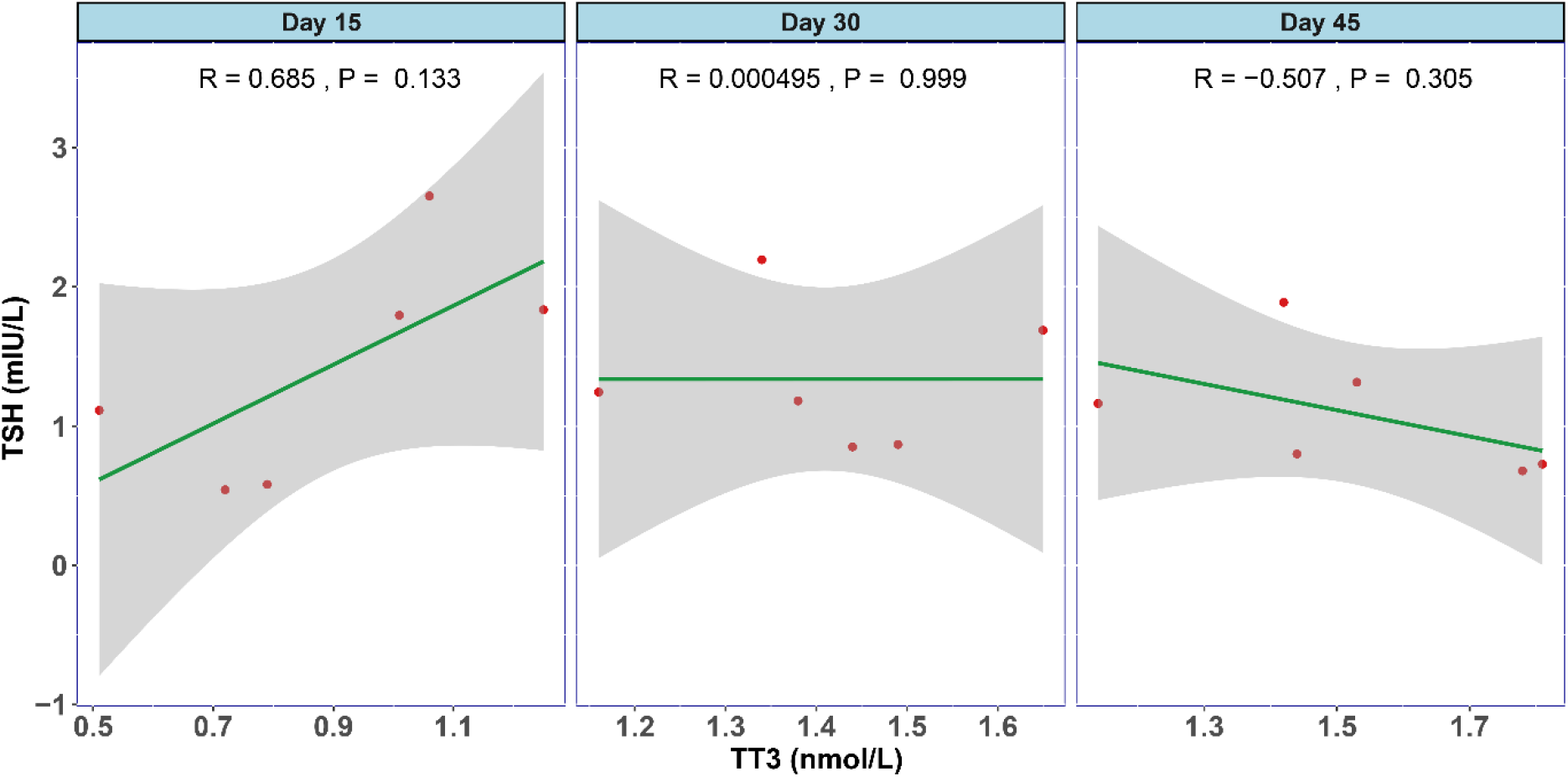
The association between TT3 and TSH levels in different disease stages of COVID-19. The levels of TT3 and TSH tend to be positively correlated in the early stage (Day 15) and turn towards negatively correlated in Day 45, but the p values are not significant.

## Discussion

COVID-19 is an infectious illness that has caused a pandemic worldwide. As a novel type of disease with high infectivity and mortality, the pathophysiology of COVID-19 has not been fully uncovered. A number of studies report the severe and complex effects of COVID-19 in several human organs and systems including respiratory, immune, digestive, circulatory, hepatic, renal, and hematological systems ^7^. However, whether COVID-19 affects human thyroid function remains unknown. Here we report the influence of COVID-19 on thyroid function. We found the COVID-19 patients had lower levels of TT3 and TSH than control groups. Thyroid dysfunction correlated with disease severity, decreased lymphocytes and increased CRP, and tended to be associated with interval time for negative conversion of viral nucleic acid. We also found that thyroid dysfunction in COVID-19 patients may recover without thyroid hormone replacement within 30 days.

Thyroid hormones are tightly related to pituitary function by TSH, and regulated by the negative feedback of pituitary-thyroid axis. Here we found that the negative feedback had disappeared in COVID-19 patients. Studies from SARS reported that the serum level of TSH and some other hormones were lower than normal range in SARS patients ^12^. Severe pathologic injury in follicular epithelial cells with follicular distortion and collapse was found in thyroid glands of SARS patients ^13^. After investigating the endocrine cells in the pituitary gland of five SARS patients, Wei et al. found that TSH positive cells were significantly decreased ^14^. These findings suggested that endocrine cells of the adenohypophysis may be attacked and damaged by coronavirus. This may partially explain why secretion of TSH cannot be stimulated by the negative feedback loop under the circumstance of reduced T3 and T4.

The underlying mechanism of damaged thyroid and pituitary function is still uncertain, which may be multifactorial and complex. Direct cytopathic effects induced by virus is a major factor ^15,16^. The wide distribution of COVID-19 nucleic acid in respiratory tract, saliva, feces, and breastmilk indicates that direct viral attack to the target cells may be an important reason ^11,17-19^. Angiotensin-converting enzyme 2 (ACE2) is a receptor providing the main entry site for SARS-CoV to invade human cells, which facilitates direct damage of virus through the course of infection ^20,21^. Li et al. recently reported that ACE2 was highly expressed in thyroid ^22^, suggesting that the thyroid gland was a potential target for direct attack of COVID-19. Our study showed that thyroid dysfunction tended to be associated with interval time for negative conversion of viral nucleic acid, indicating virus infection and replication may account for the decreased T3 and TSH. Meanwhile, TT3 and TSH levels on admission were positively correlated, suggesting that the viral damage to both thyroid and pituitary glands may be similar.

Indirect injury, such as immune mediated damage of the severe inflammatory response, also takes part in the pathogenesis of COVID-19 ^8^. ARDS is the main cause of mortality of COVID-19 ^4,6^. The host immune hyperactivity is also regarded as a pathogenesis of COVID-19 ^23^. Accumulating evidence indicates that inflammatory storm is associated with pulmonary damage in COVID-19 patients ^4,8^. A profile of cytokines, such as IL-2, IL-6, IL-7, INF-γ, and TNF-α, is associated with disease severity and may cause the mortality of COVID-19 patients ^4,6,8,16^. Our results showed that thyroid dysfunction was associated with increased inflammation biomarkers including CRP and leucocytes, indicating inflammatory reaction played an important role in thyroid impairment. We also found that the decreased lymphocyte level was associated with hypothyroidism. Since SARS-CoV has been reported to infect immune cells, mainly T lymphocytes and macrophages ^24^, we suggest that the extent of decreased lymphocyte levels may reflect the severity of viral attack and indicate the outcome of COVID-19 patients.

In the present study, we also noticed 7 patients, who had lower than normal levels of TSH and TT3 on admission, had exhibited a gradual increase of both hormones which normalized at Day 30 post-admission. Furthermore, the negative feedback of TT3 and TSH level seemed to be lost at the early stage and returned to work over time, indicating a recovery of the pituitary-thyroid axis abnormalities as well. A recent case of thyroiditis after SARS-CoV-2 infection came up by Brancatella et al. confirmed our finding that the thyroid dysfunction followed a triphasic course including thyrotoxicosis, hypothyroidism, and euthyroidism ^9^.

Since no specific and effective pharmaceutical intervention is available at present, the main treatment of COVID-19 is symptomatic and supportive. The use of glucocorticoids is still controversial and should be given at right time with appropriate dose and course ^7^. Besides, the side effects of glucocorticoids, such as hyperglycemia, hypertension, and immune suppression, should be closely monitored ^7^. Glucocorticoids can also suppress the function of adenohypophysis by pituitary-endocrine axis to inhibit the release of TSH. Considering our results, glucocorticoids should be used carefully in patients exhibiting a trend of hypothyroidism.

There are several limitations which might cause potential bias. Firstly, the study is retrospective and single centered, with limited sample size. More patients from multiple centers should be analyzed in future. Secondly, since little attention has been paid during the treatment of COVID-19, only 22 patients have complete thyroid function including FT3 and FT4. Lastly, COVID-19 patients were admitted to hospital at different stages and most of them were not dynamically monitored in thyroid function.

## Conclusions

In the current study, we found that thyroid function abnormalities were common in COVID-19 patients, especially in severe cases. The thyroid dysfunction caused by SARS-CoV-2 seems to dynamically change within the course of disease and recover gradually. Therefore, more attention should be paid to thyroid function during treatment of COVID-19, and close follow-up is also needed after discharge.

## Data Availability

Data that support the findings of this report are available from the corresponding author upon request.

## Acknowledgement

We thank Dr. Joan Ai Hui Lim from Sengkang General Hospital (Singapore) for the language editing.

## Contributors

W-WB, S-XY, D-YF, and F-WN designed the study, analyzed the data and wrote the paper. S-JW, C-ZD, ZH, X-KJ, NQ, X-XW collected data and performed the study. T-LS and Q-YQ designed the study, supervised the whole process and critically revised the manuscript.

---

**Figure S1.**
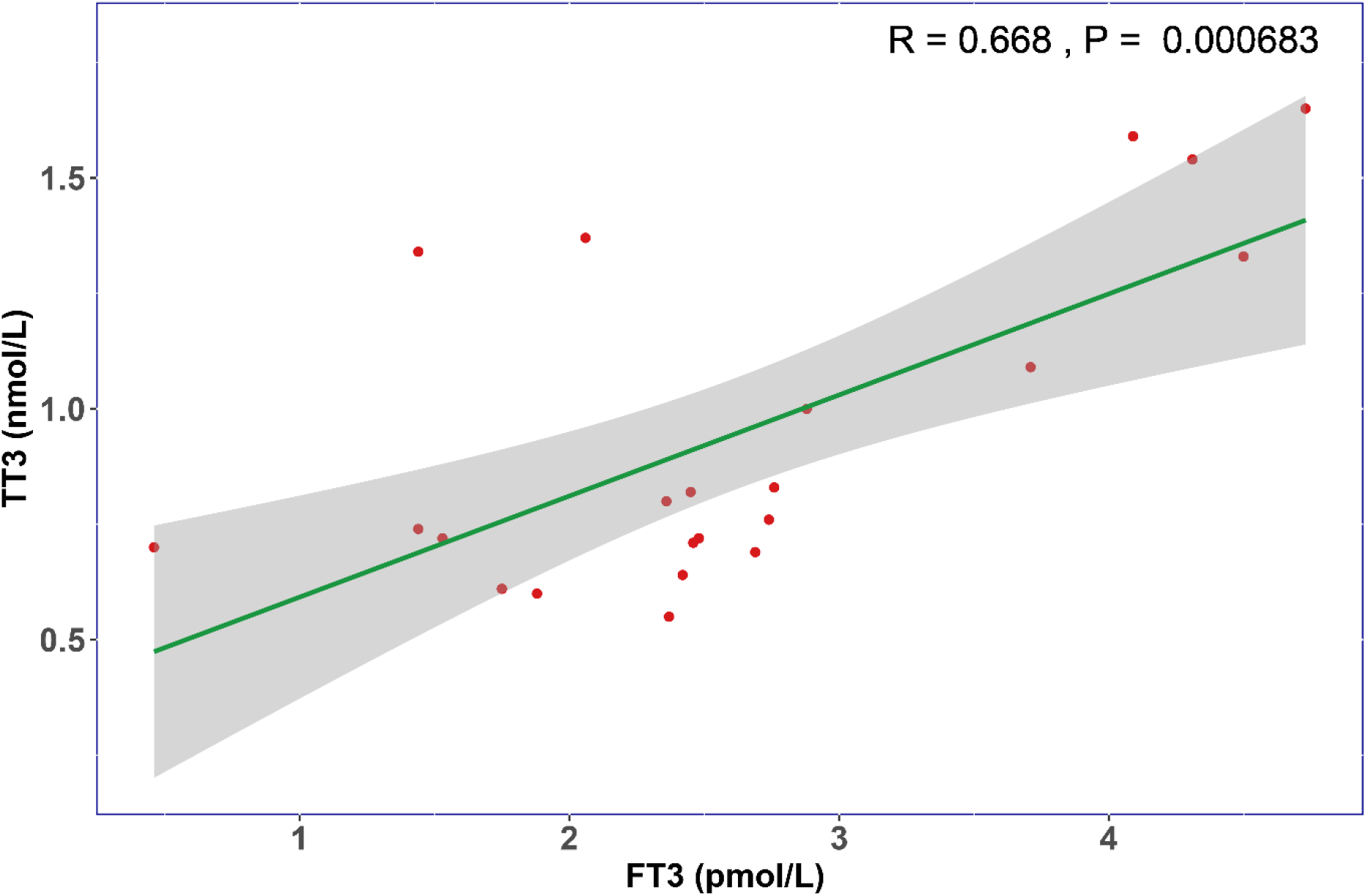
The respective association between FT3 and TT3 levels in COVID-19 patients on admission. Correlation analysis shows that FT3 is positively related with TT3 (R=0.668, P<0.01).

